# ‘We are not the virus’ – Experiences of racism among East & Southeast Asian heritage young people in London during the height of the COVID-19 pandemic

**DOI:** 10.1101/2023.10.25.23289986

**Authors:** Lu Gram, Ada Mau

**Author notes:** Corresponding author *Address:* UCL Institute for Global Health, 30 Guilford Street, London, WC1N 1EH, United Kingdom, *Email:. Joint first authors.

## Abstract

The spread of COVID-19 was accompanied by news reports of surging racism, xenophobia, and hate crime all over the Global North targeting individuals of East and Southeast Asian (ESEA) descent. However, little empirical research has documented the impacts of COVID-19 on child and adolescent ESEAs. We describe and analyse the mental health experiences of young ESEA Londoners during the height of the COVID-19 pandemic. We purposively recruited 23 young people (aged 5-24) of ESEA heritage through social media and existing ESEA networks and analysed transcripts using thematic analysis. Participants experienced distress from being exposed to multiple forms of racism ranging from strangers on the street avoiding or harassing them to classmates at school or university making racist ‘jokes’, comments or ‘banter’. Participants worried about hate crimes reported in news media and experienced anxiety from seeing pervasive racist content in online social media. Some participants responded by physically isolating themselves at home for long periods, whilst others chose to participate in activism, providing a sense of agency. Action by parents and school authorities was reported to help prevent further bullying, but respondents did not always feel able to approach these for help. Our findings put into focus the strain on young ESEA Londoners’ mental health caused by COVID-related racism and jar against simplified depictions of metropolitan places, such as London, as centres of cosmopolitanism and tolerance. To promote the emotional wellbeing of young ESEAs, future policy should facilitate action by schools and universities against anti-ESEA racism and support ESEA community-building efforts to enhance resilience in the face of racism.

---

> Interviewer: Can you share, like, a few thoughts you have about how the whole pandemic has impacted you or your family or your life?
>
> Evie: Racism.
>
> Interviewer: Right, of course. You mentioned that earlier, yes. How much did it affect you, then?
>
> Evie: Quite a lot.
>
> (Evie, aged 5-9)

## Introduction

In May 2020, United Nations Secretary-General António Guterres warned of a “tsunami of hate and xenophobia, scapegoating and scaremongering around the world” unleashed by the COVID-19 pandemic (1). News stories widely reported a surge in racism, xenophobia, and hate crime targeting individuals of East and Southeast Asian (ESEA) descent (2, 3, 4, 5). Racist, sinophobic content proliferated on social media, boosted by populist rhetoric from political leaders such as Donald Trump calling COVID-19 the ‘China virus’ or ‘Kung Flu’ (6). Even individuals racialized as ‘Chinese’ by virtue of their physical features—such as Northeast Indians living in India—were targeted and blamed (7). Three years on from the initial outbreak, activists, policymakers, and researchers are still searching for approaches to conceptualize and understand COVID-related racism.

Racism, discrimination and xenophobia are fundamental human rights and public health concerns facing societies across the world (8). Discrimination harms individuals’ physical and mental health, not only through the activation of bodily stress responses causing epigenetic change, but also through psychological responses involving anxiety, depression, and even suicide (9). Chronic exposure to discrimination and prejudice engenders internalised racism (10), stereotype threat (11), and minority stress (12), while acute stressors such as racially motivated assault cause physical and emotional injury (13). Anxiety provoked by past experiences of racism may also cause social isolation and further exacerbate mental disorders (13). Children and adolescents are particularly vulnerable, as racism can affect their socio-emotional development with long-term implications for adult mental health (14). Indeed, a prospective cohort study in the UK found that long-term psychological distress caused by experiences of racism partly accounted for inequalities in mental health between White British and minoritized groups (15).

Nonetheless, coverage of research on racism and health has been uneven across the world. A systematic review found that 81% of studies on racism and health had been done in the United States, no studies had been done in the Global South, and even within the US context, Asian Americans only accounted for 9% of study participants (16). In the UK, ESEAs are even less visible in research and policy. While the term ‘Asian’ prototypically evokes ESEAs in the US, it largely refers to South Asians in the UK, partly due to Britain’s colonial history (17). The UK census awkwardly disaggregates ‘Asians’ into ‘Indians’, ‘Pakistanis’, ‘Bangladeshis’, or ‘Chinese’ people with an ‘Other’ category for all others, whether they are Japanese, Nepali or Kyrgyz (18). Public health researchers have excluded ethnicities who were neither Black nor South Asian from large-scale longitudinal surveys on racism and health on the grounds that they were not one of the ‘main’ minority groups in the UK (19).

ESEA communities—estimated to number between 400,000 and 1.2 million in the UK (20)—have long occupied an ambiguous position in British society. On the one hand, ESEAs, particularly British Chinese, have been seen as a ‘model minority’ that disproportionately excels in education and professional life (21). This stereotype has often resulted in ESEAs being absent in debates on racism and fuelled speculations that ESEAs do not experience racism (22), despite evidence to the contrary (23). On the other hand, ESEAs have been cast as immigrants, take-away workers, and triad gangsters (24), which has historical links to structural violence—in 1945, up to 20,000 Chinese seamen in Liverpool were forcibly repatriated by the UK government in spite of having committed no crime (25).

COVID-19 starkly demonstrated the precarity of ESEA status, when ESEAs were abruptly cast as foreign disease carriers (26, 27). An event study of UK police data concluded that racial hate crimes against ESEAs increased by 70-100% at the beginning of the pandemic in early February and persisted until November 2020 (28). In educational settings, the sudden surge of racism prompted the teachers’ union, NASUWT, to urge the then Education Secretary, Gavin Williamson, to address COVID-related racism in schools (29). All this emerged at a time in which children and adolescents faced many mental health challenges due to COVID-19 in general, including exacerbation of existing mental health conditions (30). However, only a handful of empirical studies provide evidence on the experiences of COVID-19 on ESEAs living in the UK, all of them involving primarily adult participants (31, 32, 33). As such, there is need for evidence on the impacts of COVID-19 on ESEA children and adolescents. We describe and analyse the mental health experiences of young ESEA Londoners during the height of the COVID-19 pandemic.

## Methods

We applied qualitative research methods to explore the lived experience of young Londoners of ESEA heritage. Our overall approach was informed by phenomenological research, which aims to study how people experience a shared phenomenon (in this case the COVID-19 pandemic) (34). We report methods in accordance with COREQ guidelines for qualitative research (35). AM and LG are both UK-based middle-class academics of ESEA heritage with extensive experience in qualitative research. As ESEA people ourselves, we had a genuine interest in the research topic, hoping to find ways to support the ESEA community. AM has conducted research in British Chinese communities previously (36).

### Sampling and recruitment

We employed snowball sampling, purposively recruiting ESEA participants through social media and existing networks, including support from external partner organisation End Violence and Racism against ESEA Communities (EVR-ESEA) (37). All participants of ESEA heritage, aged 5-24 years’ old, and residing in London were eligible. We established a website for recruitment (38) and advertised our study on Instagram, Facebook, and Twitter. The call for participants generated tremendous support from ESEA networks, communities, and well-known public figures such as actor Gemma Chan and singer Emmy the Great, indicating that people wanted to be heard and to raise the profile of this issue. As part of this process, AM had access to information identifying participants, so that she could contact them for recruitment.

We conducted semi-structured interviews with 23 young people aged 5-24 living in London of East and Southeast Asian background from May to August 2021. Table 1 provides demographic information individual respondents. The participants came from different regions of London, both inner and outer. 13 were Chinese, three Vietnamese, one Malaysian, one Filipino, and five of mixed heritage. They were all in education, with the exception of two who were in a gap year before starting university. We categorised participants’ social class based on their parents’ occupations and other relevant information they provided in the interview. The majority of the young people (70%) came from middle-class households.

**Table 1.**
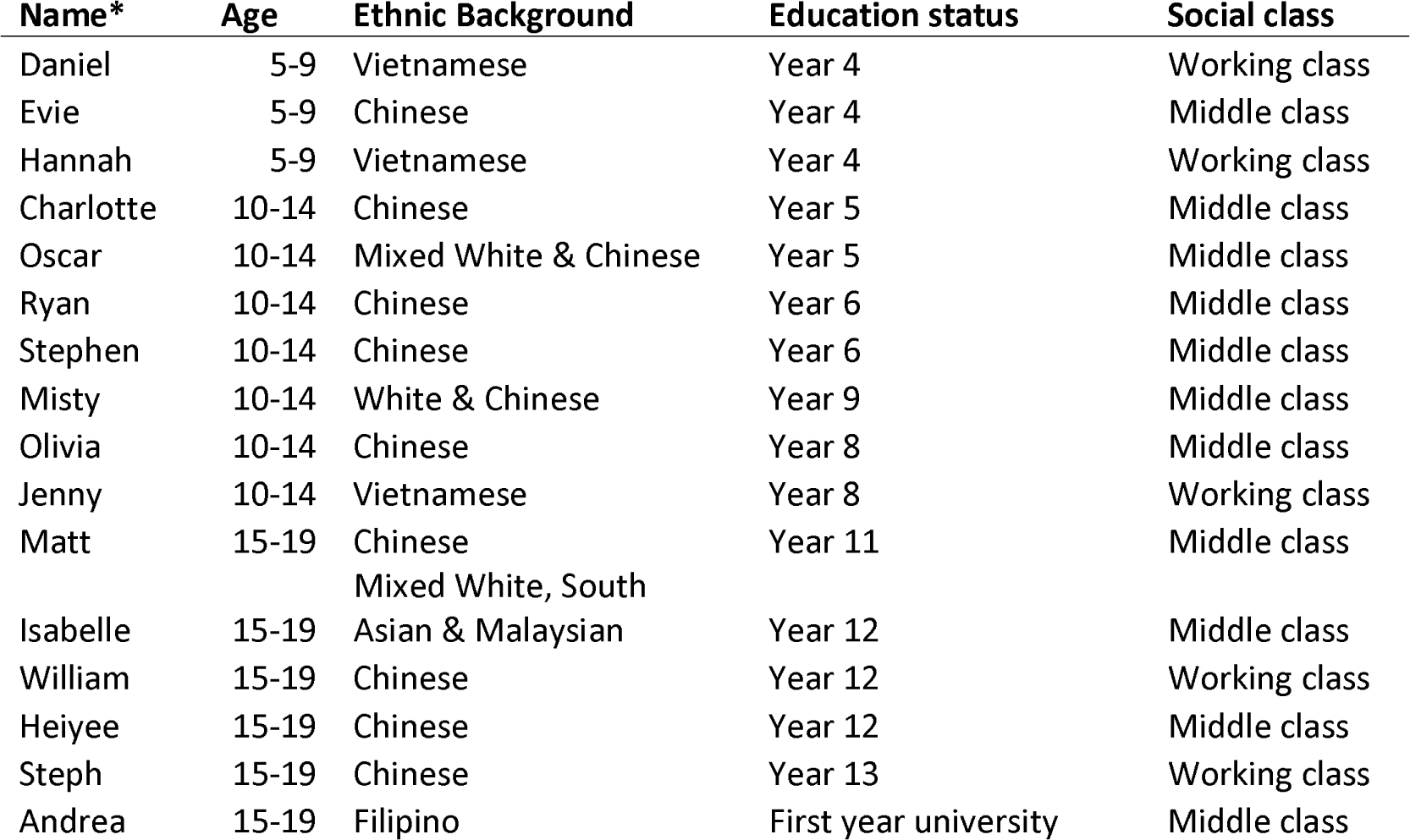
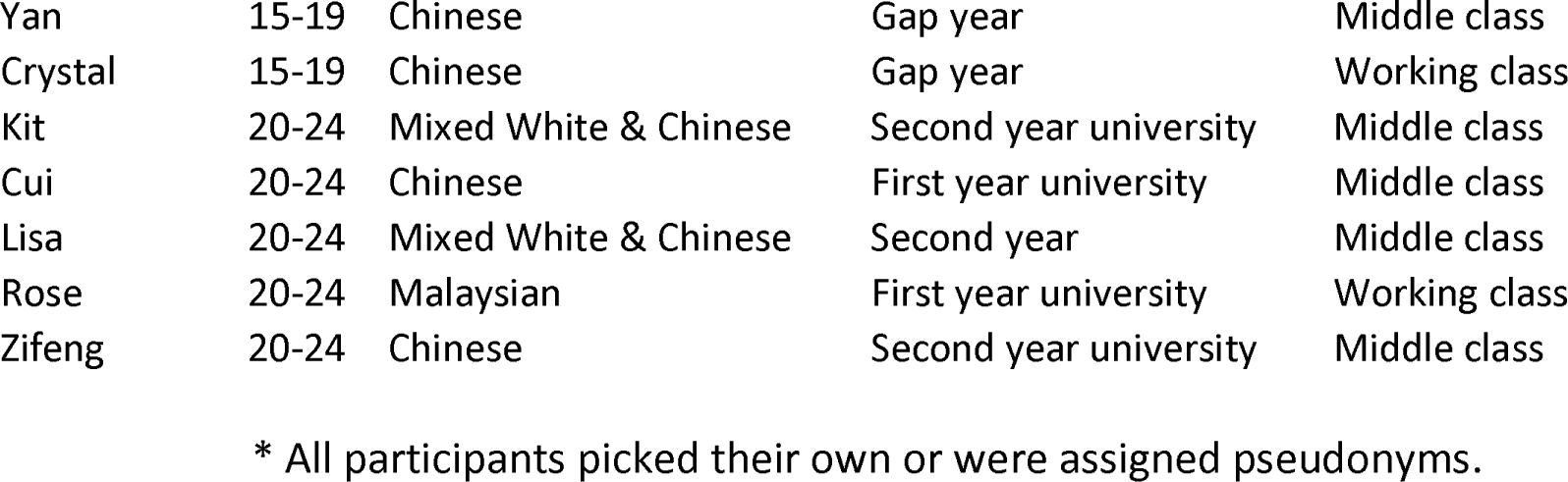
Demographic information of participants.

### Data collection

We developed and piloted a topic guide for semi-structured interviews covering young people’s lived experiences on education, health and wellbeing, living environment, including questions on mental wellbeing. Author AM conducted all interviews online in English. Neither author knew any of the participants prior to the interview except for one participant whom AM knew through a personal contact. None refused to participate. Due to pandemic restrictions, interviews were conducted on recorded video or voice calls. In case of younger children aged under 12, parents were also present, but did not speak during the call. Participants received a £10 voucher as compensation for taking part. AM wrote personal reflection notes after each interview. Interviews were transcribed using a combination of manual correction to automatic transcriptions done by Zoom and professional transcription by paid transcribers.

### Data analysis

We conducted a thematic analysis following Braun and Clarke (39). We followed all six steps of their method, including familiarisation with data, generation of initial codes, search for themes, review of themes, definition and naming of themes, and production of the final report. We used both inductive and deductive coding, approaching the data with a view to analysing the data using the concepts of racism, mental health, and social identity, while remaining open to participants’ own understanding of their lived experience. Author AM familiarised herself with all transcripts and coded them, while author LG coded three transcripts (13%) and compared the resultant codes with AM to strengthen rigour and conceptual clarity. AM and LG discussed codes identified to increase reflection over the data. Coding was conducted in NVivo. We identified major themes concerning disruptions to daily life, COVID-19 related racism and responses to such racism by respondents. The full coding tree with example quotes has been provided as Supporting Information.

### Ethics

This study was approved by the University College London (UCL) research ethics committee (REC1483). The purpose of the research was explained to study participants and informed consent was sought prior to and during the interview. Written informed consent was obtained from the parent/guardian of each participant under 18 years of age.

## Results

### Disruption to daily life

Respondent accounts revealed that the height of the COVID-19 pandemic in the UK was a difficult period. The changes to normal life and routines during lockdowns generated frustration, as participants felt stuck in a lockdown bubble. Haiyee (aged 15-19) had even ended up changing her sleep cycle, so she was awake during the night and slept during the day for three months. Further, although most managed remote learning the best they could, they admitted it was exhausting due to lack of in-person interaction and unsettling for those facing exams or about to start university due to the poor quality of learning. Lisa (aged 19-24) was starting university life remotely felt like ‘*a time of my life that I’m supposed to cherish forever, and it’s supposed to be some of my best memories are going to waste.*’

Some got very worried about themselves, or their family members being infected by this new and unknown virus. For example, Daniel (aged 5-9), whose father ran a restaurant, felt ‘*really worried*’ about himself and people around him getting ill, reporting that he ‘*got ill because of that* [his worries]’. Although many respondents had parents who were in professional jobs who could work from home, some had parents who had to continue to travel to work and face the public, reflecting the fact that a substantial proportion of UK ESEAs are employed in the catering and beauty trade, which suffered considerably during the pandemic. Others had parents working in healthcare who were regularly in contact with patients suffering from COVID-19.

### COVID-related racism

#### Street encounters

ESEA young people expressed distress at being subject to uncomfortable and sometimes aggressive street encounters making them feel vulnerable and unwelcome. A number of participants spoke about people seemingly avoiding them or their family members and friends on the street or on public transport. For example, Isabelle (aged 15-19) noticed that ‘*people would cross the road to avoid you because of your look*’. Lisa (aged 20-24) described a hostile encounter on a train with her mother and grandmother, in which someone ‘*came next to us and looked at us and opened the window next to us and then moved and sat in a different seat.*’ A number of participants also shared stories of family members or friends being followed. Matt (aged 15-19) shared an unsettling experience of his mother being surrounded and circled by cars in the street:

> One time we were walking my dog, with my mum, my older brother, my little brother, and my cousin, so, a lot of people. My mum was ahead… walking ahead, and my mum found that there were people in cars circling and looking at her and then they left when all of us, all of our brothers and cousins caught up. I guess that’s when we first started seeing it in real life.

#### ‘Jokes’, remarks, ‘banter’

Respondents often reported experiencing racism in the guise of ‘jokes’, causal remarks, or ‘banter’. Participants spoke about remarks, such as ‘Kung Flu’ and ‘China Virus’, being said in schools, universities and in wider society. Some of the other racist remarks were built on the pre-existing prejudice against ESEA cultures and particularly food cultures. For example, a university student, Cui (aged 20-24) described hearing ‘*bat soup*’ from his course mates, including in a comment made in front of the whole a class. Rose (aged 20-24) discussed how, at the start of the pandemic, her course mates made negative comments about other East Asian people wearing masks.

Younger children were also targeted, as we heard from participants across all levels of educational settings, from primary school to university, reported racist encounters. Daniel (aged 5-9) described people saying ‘*really rude things*’ relating to the virus being from China and about Chinese people, even though he could not clearly remember or did not want to articulate the exact things were said when being asked. Charlotte (aged 10-14) also talked about pupils at her school saying things she ‘*didn’t really like*’ about the virus and would point at her and her ESEA friend. Importantly, perpetrators of racism often included members of other minoritized groups. As Matt (aged 15-19) explained: ‘*The majority of the school were doing it so, brown, Black. I guess the majority of people in my school are brown, so I noticed a lot more from brown people.’*

#### Online racism and news media

Participants were often indirectly affected by racism through awareness of – and subsequent worry about – serious incidents of physical violence reported in news media. Many of the participants were highly aware of the development of anti-Asian American racism in the US. The high-profile violent, racist attacks in the UK on university student Jonathan Mok and lecturer Peng Wang (40, 41) shocked many in the ESEA community and even young participants, such as Matt (aged 15-19) were aware and expressed worry over UK hate crime statistics: ‘*We were concerned about safety, from the rises of Asian hate crimes. My mum was quite worried about that, and I was too. I didn’t want anything to happen to my parents … the statistics were really worrying’*

Racist online comments also affected participants, even if they were not directed at them personally, as they contributed to a hostile backdrop experienced regularly. For example, Heiyee (aged 15-19) recalled a Chinese New Year’s greetings post by then Prime Minister Boris Johnson receiving many racist comments online, blaming Chinese people for COVID and the lockdown. Many participants mentioned seeing comments about Chinese people eating bats or dogs online. Kit (aged 20-24) reported regularly seeing casual racist comments online, then hearing them offline from people around her, such as, ‘*Oh, I wish that Chinese guy just hadn’t eaten that bat!*’ Although she felt things had improved after the end of lockdown, she had only to glance at social media to confirm the continuing existence of anti-Asian sentiment in British society.

### Responses to COVID-19 racism

#### Physical isolation against street harassment

A number of young people spoke about isolating themselves at home due to the combined worries of contracting the virus and receiving racist abuse. In fact, Zifeng (aged 20-24) felt so vulnerable at her hometown outside of London, a predominantly white area, and did not leave the house for 2.5 months because of the fear of racist attack. Rose (aged 20-24) talked about being ‘*always on my guard now*’, feeling resigned to a state of wariness, ‘*I think that’s really sad, isn’t it, to have to feel that way when you’re going out.’* Respondents also worried about the safety of other family members, whilst simultaneously finding the topic difficult to discuss openly. For example, Kit (aged 20-24) struggled to voice her apprehension at her mother talking regular long walks in the face of COVID-19-related hate crimes:

> I told her to just, I was like, “You know what’s going on, right?” And she was like, “Yeah, I know.” I was like, “You should stay…” I don’t know, I found that very uncomfortable, I didn’t know what to say … but I was just like, “Just stay safe, it’s pretty weird out there.”

#### Struggling to react to ‘jokes’ and casual remarks

It was often difficult to deal with casual remarks made by known others inside and outside of educational settings. Young people spoke about ‘everyday’ experiences prior to COVID-19 of racist name-calling or gestures (people making ‘Ching Chong’ sounds or squinty eyes) and subtle forms of microaggression (stereotyping ESEA pupils as ‘model students’, pathologizing ESEA food cultures) that they had received little help with from teachers. Thus, some participants tried to ignore COVID-related racism, as they did with previous incidents. For example, Steph (aged 15-19) talked about how she was too shy to confront perpetrators and would ‘*just laugh it off and just move on*’, only later on even realising ‘*how much prejudice and racism was labelled as joking or banter’.* Others—such as Kit (aged 20-24)—tried to engage speakers but found themselves arguing with them:

> …just in a casual conversation, “Oh, I’m going to Chinatown after this to get some food.” And he was like, “Well, you know Chinese people eat cats and dogs,” and I was like, “Well, you know, I don’t think that’s true.” And he was like, “No, no, they do, they actually do.” And I was like, “I don’t even know what to say to this.” But it was so uncomfortable, and it was also the height of summer, and I was just very like, “I don’t want to try and argue with you.”

#### Helpful actions by parents and school authorities

Swift action by parents and school authorities was often reported to be helpful. Evie (aged 5-9) was called a ‘*monster*’ by her classmates due to her ethnicity and her brother Ryan (aged 10-14) recalled that ‘*someone came up to me and started blaming me for the coronavirus*’ at school. Their parents reported the incidents, their school dealt with the problems swifty, and the abuse stopped for them. Isabelle (aged 15-19) was told by a classmate that he did not want to sit near her in the cafeteria, as he was ‘*going to catch corona’.* In response, she coughed on him as revenge and reported him to school authorities who called his parents for a conversation and suspended him.

However, respondents did not always report their experiences to parents or school authorities, due to communication barriers with parents, disbelief in the ability or willingness of teachers to change peer norms, or prior experiences of inaction by teachers: ‘*Unless they were specific incidents, most of it was just random comments from random people and you can’t exactly make a huge fuss out of that!*’ (Misty, aged 10-14). The experience of racism also cast a long shadow. Evie (aged 5-9) was still visibly upset during the interview when discussing her experience of being called a ‘monster’ and the interviewer had to stop the interview to let her collect herself before continuing. When asked to summarize the impact of the pandemic on herself, her family, or her life, she simply said, ‘*Racism.’*

#### Participation in anti-racist activism

Some respondents—particularly older ones—expressed an awareness of the role of broader societal factors such as media or social norms in driving racism against ESEAs. For example, Kit (aged 20-24) described the inflammatory role of media reporting on COVID-19:

> The way that the media … continues to report on anything sounding like Covid, definitely contributes to it [racism] so much. It’s just all these really subtle things, or not so subtle, really, just like the types of pictures that they choose, the types of really inflammatory headlines that they choose to sell their papers and everything. A lot of people blame the media for lots of things but, in this case, I really do feel like it was blown up by them.

Simultaneously, respondents discussed the rise of the Stop Asian Hate movement, anti-Asian hate crimes in the US, and the Black Lives Matter protests, which prompted some to engage with their own communities. Williams (aged 15-19) reflected on how the conjunction of COVID-19-related racism and Stop Asian Hate led him to think much more frequently than before about his identity as British and Chinese. Heiyee (aged 15-19) described how she had greater empathy for the experiences and struggles of Black people, *“I finally understood what it felt like to see people who looked exactly like me be hurt in a way that I could never imagine because of something that we couldn’t control.”*

As a result, some of the participants turned to activism to help combat the rise of anti-ESEA sentiment. For example, Kit and Zifeng (both aged 20-24) became actively involved in online activism aiming to empower British Asian young people. Matt (aged 15-19) started doing graphics for an Instagram account that supported ESEA people in the UK. It made him feel like he had greater agency over a phenomenon that was otherwise outside his control:

> Like, you look at some of the stuff that happens online, and you feel useless and hopeless because you couldn’t do anything, watching a video, but, when you’re actually doing something, it makes you feel a bit better. It makes you feel less useless. It might not be much, but it might be helping some people.

## Discussion

In addition to laying bare ethnic health inequalities in the UK (42), COVID-19 plainly demonstrated a need for research into the mental health of ESEAs as a group. We found that young ESEA Londoners’ emotional wellbeing—already worsened by social isolation, school/university closures, national lockdowns, and worries over parents’ livelihoods—faced unprecedented pressure from COVID-related racism in 2020 (Table 2). Young ESEAs were widely exposed to microaggressions, bullying, and racist comments—both online and offline. ESEAs not directly exposed to racist remarks experienced anxiety over a generally hostile environment and reported increases in hate crimes in police statistics to the extent that some avoided leaving the house for long periods of time for fear of racist attacks. Support from education authorities and parents were helpful in addressing bullying and racist comments, but young ESEAs did not always feel able to seek help. Turning to activism helped young ESEAs reclaim a sense of agency over otherwise uncontrollable events.

**Table 2.**
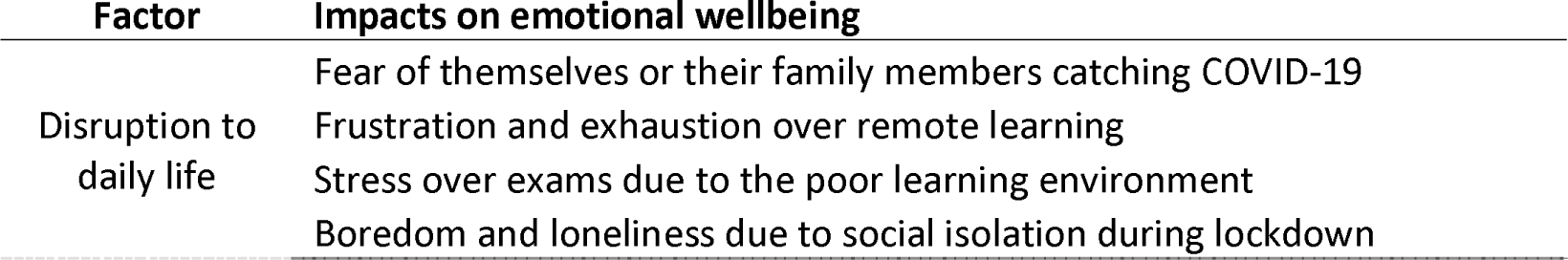
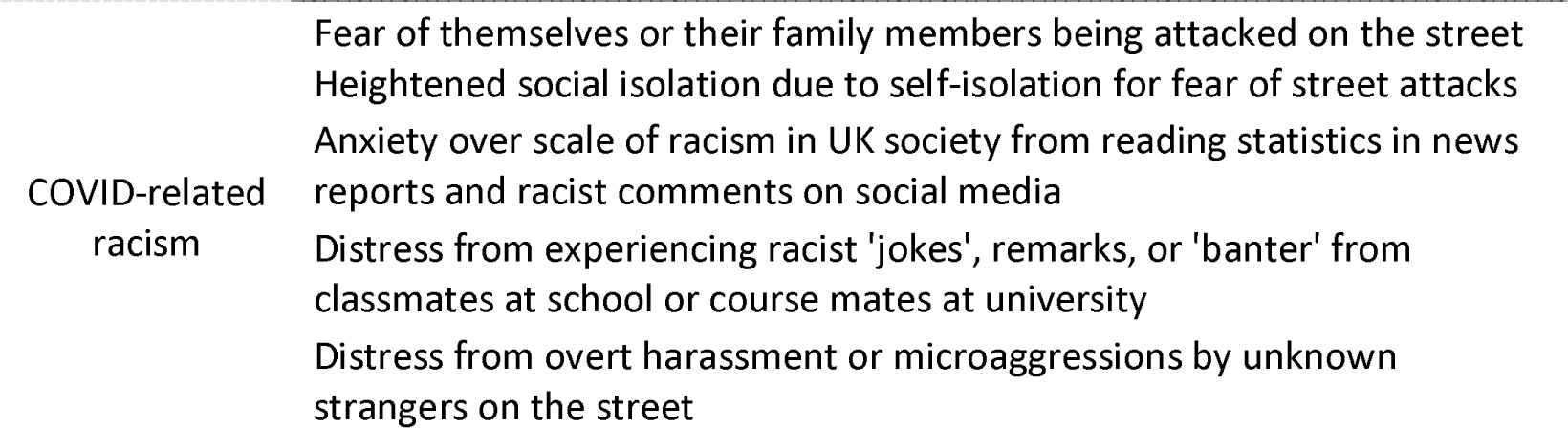
Impacts of COVID-19 on the emotional wellbeing of young ESEAs.

| Factor | Impacts on emotional wellbeing |
| --- | --- |
| Disruption to daily life | Fear of themselves or their family members catching COVID-19 |
|  | Frustration and exhaustion over remote learning |
|  | Stress over exams due to the poor learning environment |
|  | Boredom and loneliness due to social isolation during lockdown |
| COVID-related racism | Fear of themselves or their family members being attacked on the street<br>Heightened social isolation due to self-isolation for fear of street attacks<br>Anxiety over scale of racism in UK society from reading statistics in news reports and racist comments on social media<br>Distress from experiencing racist 'jokes', remarks, or 'banter' from classmates at school or course mates at university<br>Distress from overt harassment or microaggressions by unknown strangers on the street |

Much existing discourse on COVID-related racism has focussed on hate crimes against ESEAs, which increased dramatically during the height of the pandemic (28, 43). Our findings highlight the need to consider alternative framings of COVID-related racism, as many of our respondents experienced racism in the form of racist remarks made by people, they knew—classmates and friends, some of whom are also from ethnic minority backgrounds. They or their parents were unlikely to report friends or classmates to the police for committing a ‘hate crime’. Many respondents also experienced COVID-related racism as a continuation of ‘everyday racism’ (44) prior to COVID-19, bringing up common past experiences (people making ‘Ching Chong’ sounds or squinty eyes). Rather than a passing crime wave, the emergence of COVID-related racism arguably reflected an intensification of existing Orientalist prejudices in British society; COVID-19 may have served as a vehicle for voicing and transmitting those biases (23, 25).

Our findings jar against depictions of metropolitan cities, such as London, as ‘centers of cosmopolitanism, that is, as places in which people have access to multiple lifestyles, confront diversity on a daily basis, and frequently adopt a tolerant attitude that accepts social and ethnic differences with a minimum of rancor’ (45). Past debates over urban multiculturalism in the UK have focussed on the politics of migration, integration, and racial segregation in the wake of the US War on Terror (46, 47, 48). We argue that these discussions must also (re)assess multiculturalism and diversity in light of ESEA experiences. The fact that many perpetrators of racism in our data were non-White young people challenges the thesis that large, multi-ethnic cities such London are places in which cultures and ethnicities mix ‘with a minimum of rancor’ (45). It also highlights the need for approaches to anti-racism which foster solidarity between minoritised groups and addresses multiple forms of racism simultaneously (49).

A few young ESEAs in our study had indeed participated in anti-racist volunteer networks to address COVID-related racism. Multiple such networks emerged in 2020 and 2021 (50), some of which now provide mental health and counselling services for ESEAs affected by racism (51). Our findings highlight the different faces of online media, both as sources of disinformation and racism and as places of community and sharing of difficult experiences (52, 53, 54). Indeed, the very idea of a pan-ESEA identity in the UK initially emerged as a social identity to support the mobilisation of political community (54). These forms of community building may mitigate the damage wrought by COVID-19 and contribute to community mental health competencies (55).

However, the onus on addressing racism and its harms should arguably not lie on those who suffer from racism themselves. Education researchers have documented sinophobic racism—often banal and subtle, yet harmful—affecting ESEA young people in UK schools for decades (56, 57). To promote the emotional wellbeing of young ESEAs, schools need to develop a whole-school anti-racist culture in supporting children of all backgrounds. This involves having ongoing discussions on racism in the school community, so as to be adequately prepared for large-scale socio-political developments such as COVID-19. An example of such an approach is the Anti-Racist School Award framework (58), which evaluates the degree to which anti-racism is embedded in the entire school system (from governance to pedagogy) and internalised by all staff (from admin to cleaning staff).

### Limitations

Due to pandemic restrictions, we were unable to recruit in-person at community events or activities. Therefore, the sample had more middle-class participants, who might be more connected online and more forthcoming in participating. Although our call for participants did not explicitly mention COVID-related racism (38), our external partner organisation EVR-ESEA focuses on addressing racism against ESEAs (37). As such, we may have sampled more ESEAs with experience of racism or who were attuned to discourses on racism; our findings should not be assumed to represent all ESEAs. It is also worth noting that not all of our respondents spoke of experiences of racism, indicating a diversity of ESEA experiences during COVID-19. Our phenomenological approach provided rich data on ESEA lived experience but did not always provide detailed statements on causes of distress—as our respondents might be unaware of the underlying causes of low mood or face difficulty in articulating those causes. Finally, the cross-sectional nature of our sample meant that our study focused on ESEA experiences during the height of the pandemic, including lockdown.

### Future directions

We found that COVID-19 unleashed widespread racism on young ESEA Londoners that they themselves, their parents, and schools struggled to respond to. Our findings show that public health researchers and practitioners, including mental health researchers, working in contexts with ESEA minorities must take into account the differential needs and experiences of ESEAs. Our findings show that the mere fact that educational institutions are situated in a metropolitan setting such as London does not exempt these places from harbouring racism. Prejudice-reduction, anti-racist interventions in educational settings (59) may be an important priority for future research. Together with greater support for ESEA communities and wider reform of school and university policies, we can move towards more inclusive environments that enable young ESEAs to truly thrive.

## Data Availability

As this is qualitative research data, and reports on the experiences of racism of young people, we opted not to share the data publicly. We also did not obtain consent from participants to share their data in an open access fashion.

## Acknowledgements

We thank End Violence and Racism against ESEA Communities, Gemma Chan, Emmy the Great, and many others for their support with recruitment of participants. We thank the participants who generously provided time to speak about their, sometimes difficult, experiences. This work was funded by a UCL Grand Challenges Small Grant (2021).

