## Supporting Information for "‘We are not the virus’ – Experiences of racism among East & Southeast Asian heritage young people in London during the height of the COVID-19 pandemic"

Supplementary data: Thematic frameworks Appendices

A1. COVID-related racism thematic framework table

| *Thematic Category:* | **COVID-related racism** | |
| --- | --- | --- |
| *Themes* | *sub-themes* | *Sample Codes* |
| **Online and public discourse** | COVID-racism towards Chinese or ESEA people; comments blaming China/Chinese people | Even looking at some of the comments on things when the Prime Minister said: “Happy Chinese New Year,” and you see some twisted comments like: “No, no, they’re the reason we’re in lockdown. It’s because one of them ate a bat. You shouldn’t be wishing them a happy new year, I hope they have a crap new year.” *(Isabelle, aged 15-19)* |
|  |  | So I guess outside of school, on social media there was a lot of stuff going on, especially keeping up with like what political figures have said. It was really interesting. Like all Donald Trump had to say, that was something that was really … I guess it was … I wasn’t really surprised to know that was him, he said that kind of stuff. It was still because he has all the influence that he has it was really disheartening. *(William, aged 15-19)* |
| **Encounters at school/university** | Racist bullying; ‘jokes & banters’; misinformation on COVID blaming China/Chinese people | Like, for example, if you sneeze in class or something at that time, people will be like, whoa, and you know, they'd start laughing, you know, make jokes about it. I wouldn't really call it hard bullying. Like, I never got physically, you know, attacked but it was like, just passing remarks or like jokes or smirks and stuff. *(Matt, aged 15-19)* |
|  |  | There was a bit of misconception of the whole eating a bat thing. That misconception spread around the school quickly and I wasn’t sure at the time. *(Olivia, aged 10-14)* |
| **Encounters in public spaces (themselves and people they knew)** | Being avoided; racist aggression; being followed | I think a few times there. And then I do remember, when I was walking down the street, there was a guy, and he had a scarf because it was like, yes, it was about that time when it first started, February-ish. And then when I walked down, he got his scarf and wrapped it around his face, and then I turned back to look to see what he was doing because I thought, I don’t know, because I was like, you’re always second guessing yourself because you’re like, are you actually, why are you doing that? Yes, anyway, and then I looked back and he didn’t do it to anyone else, and I just thought, oh my God. *(Rose, aged 20-24)* |
|  |  | I can’t think of any on campus but my friend did say that in the supermarket it was like having an aura around him that no one wanted to go near. He said that people were literally clearing aisles to get away from him. I think there was another uni where these Chinese students in the supermarket were getting harassed by younger people and having shopping trolleys basically thrown at them, pushed in their direction and stuff. *(Lisa, aged 20-24)* |

A2. Responses to COVID-related racism thematic framework table

| *Thematic Category:* | **Responses to COVID-related racism** | |
| --- | --- | --- |
| *Themes* | *sub-themes* | *Sample Codes* |
| **Avoiding being outside** | Not going outside for extended period of time | From the first, when, after the first lockdown when restrictions were relaxed, I didn’t see anyone for three months. I think partly because I myself was scared, but also scared of going out on the street, especially hearing what was happening, what is happening with East and Southeast Asian people being attacked. I was just like, do you know what, that’s a lose/lose for me, I don’t want to get COVID and I don’t want to get beaten up. *(Rose, aged 20-24)* |
|  |  | Interviewer: Is that one of the reasons why you mentioned earlier, like you haven’t been out as much in the last few months?  William: Partially, but there’s also been like … because of how the virus has been spreading that was also part of the reason.  Like the risk of catching Covid, right, I see.  William: Yes but also because of being scared of people were being treated. *(William, aged 15-19)* |
| **Worried about their family members** | Worried about family members being vulnerable to potential racist incidents | I was really scared that they [her parents] would be attacked. See, they have like Chinese accent so they're more likely to be mocked. *(Zifeng, aged 20-24)* |
|  |  | I know my grandma likes to go out on walks on her own so that did worry me because she’s an easy target because she’s old and vulnerable. I wasn’t made to feel that way about anyone else but just because of that atmosphere there were times where I did feel quite worried but thankfully I think close to my house, I live in quite a safe community and the places where my grandma would walk, the parks would never be empty. They would always have some people dotted around so if she ever ran into any danger, there would always be the security of hopefully someone coming to help her. *(Lisa, aged 20-24)* |
| **Inability to confront racist incidents** | Ignoring racist remarks; difficulty in dealing with racism; difficulty in confronting racism not directed at them personally | Well just random strangers in the corridors, if they saw someone. Thankfully I avoided most of that because apparently I don’t look Asian enough. They were just telling people that they ate bats and they blamed them for the Coronavirus. *(Misty, aged 10-14)* |
|  |  | Interviewer: At school, was anyone making fun of the virus being from China at all?  Daniel: Yes.  Interviewer: What did they say, do you remember?  Daniel: Not sure.  Interviewer: Did they make fun of it, like, related to Chinese people?  Daniel: Yes.  Interviewer: Can you remember what they said?  Daniel: No. Not quite, no, no, nothing.  Interviewer: Okay. That’s fair enough. Do you remember how that made you feel at all, like, you hearing any of those things, if you remember?  Daniel: I felt they were really rude.  Interviewer: They were really rude? Were they saying it to you or just saying things in general?  Interviewer: Just saying things in general. *(Daniel, aged 5-9)* |
| **Activism to support ESEA communities** | Participate in activism groups and activities | I was upset about the racism towards East Asians have felt like whenever I tried to speak how people be like oh that's not a big deal, and they'd kind of downplay. And then I found this platform on Instagram and they were like they advertise something saying to begin your own chapter, I thought I’d begin one in the UK, because most of the adaptation racism just always centred on America and, like in the UK there's a lot less. . *(Zifeng, aged 20-24)* |

A3. Pre-existing racism thematic framework table

| *Thematic Category:* | **Pre-existing racism at school** | |
| --- | --- | --- |
| *Themes* | *sub-themes* | *Sample Codes* |
| **Racist ‘jokes’, ‘banters’, confrontations** | Racist name-calling, comments, gestures, violence | Interviewer: Before the coronavirus started, did you ever experience any of those kind of issues before at school, like, people making fun of you?  Ryan: Yes, definitely.  Interviewer: If you don’t mind sharing, what did people say before, like, things they said before?  Ryan: Once this other kid, he just came up to me and said that I ate slugs. And then my friend, he got a bottle of water and he sprayed it at him. I don’t know why he did that. And then there was another time when one of my old school mates who has left now, he whacked me in the eye with a ball. So then the parents came over to have a conversation with the teacher and then his brother said, like, the C-word to me.  Interviewer: That’s not nice. What about you? Were there people saying things before?  Evie: The same but someone pulled their eyes at me.  Interviewer: Sorry, somebody…?  Evie: Pulled their eyes at me. *(Ryan, aged 10-14 and Evie, aged 5-9)* |
|  |  | Yes, I’d say so, you get that a lot and people would try and imitate the language, your typical “ching chang chong” kind of thing, people would definitely say that. I suppose also the question: “Where are you from?” and then you reply: “London, Britain,” and they go: “No, where are you really from?” *(Isabelle, aged 15-19)* |
| **Microaggression** | Stereotypes, Othering | I feel like, in [x] school, I’ve been told that I mightn’t have got on this course because I’m too timid or whatever, and I appreciate feedback, but at the same time, I’m like, part of me goes, are you calling me timid because of this stereotypes around my race, or am I actually, because I feel as though if you actually get to know me, I’m not timid. *(Rose, aged 20-24)* |
|  |  | I was also throughout primary and secondary, I was very self-conscious about my accent, because people would be saying, I have a Chinese accent, it is not. It… doesn't necessarily rude, but it makes me feel uncomfortable It makes me feel like I don't speak like others, so yeah I was just always conscious about my accent not very good, I felt like I was speaking perfect English. *(Crystal, aged 15-19)* |

A4. Other pandemic related factors affecting mental health thematic framework table

| *Thematic Category:* | **Other pandemic related factors affecting mental health** | |
| --- | --- | --- |
| *Themes* | *sub-themes* | *Sample Codes* |
| **Remote learning** | Difficulties with online learning, lack of motivation; change of learning environment; worries about impact on grades/future plans | To be frank I just hate it, I still don’t like it that much. I’m so much better in class learning. I much prefer it, if I can like see the teachers face to face. I often learn better from others when I‘m with them. So, when it was all online, I just get distracted on my phone, I’m not motivated. I just find it so hard, which is probably why I didn’t do as well as I did in my exams and work, in general. I just don’t think I did that well as I could have done, considering that, I like, I did get like good grades at sixth form. It’s kind of, just, like kind of a let down because of like online than school, yes. *(Andrea, aged 15-19)* |
|  |  | It wasn’t even just the remote learning that I hated but I never felt like I wanted to do any work because it just felt so not… it just didn’t feel like school. It just felt like I was forced to go to this lecture for an hour and wake up at the same time that I normally would. I’d just be so tired all the time and I’d be so lethargic, just never wanting to move and get up because I barely left the house as well. It was always the same environment and I got so sick of it. So even when they set work, I never had the motivation to do it. *(Heiyee, aged 15-19)* |
| **Social isolation** | Loss of contact with friends; missing friends, extended family and other social contact | It was quite an emotional roller coaster for me during the pandemic at the beginning, I ended up making myself very busy because I thought, like look I have all this time on my hands I should do something worthwhile, so I did online courses. I tried to do more exam questions or spend time reading as well, and so I kind of like shut myself away from my friends. I didn't get the impact until I felt like so low at times and I would attribute that to not having enough social contact. *(Crystal, aged 15-19)* |
|  |  | Yeah I’d be missing my other family members not being able to go and see what they're doing and I would miss my friends and people, my classmates in class. *(Charlotte, aged 10)* |
| **Disruption to routines and normality** | Loss of normal routines; blurring of home/work time and spaces; changes in local environments | It just felt like there was nothing I needed to do outside. There was nothing I needed to do at home either but then I just kind of flocked to the option where the least effort was required because if I had to go outside then obviously I had to put clothes on, probably take a shower or something, really force myself to move my body. *(Heiyee, aged 15-19)* |
|  |  | It was definitely less, there were less places open. There were still some, a little bit, but definitely not as much. That was really sad. And then seeing some of our favourite places closed down as well, some places that we’d gone to quite a bit, that was really sad. Because we’d talked to the people who had worked there, and things like that, so it was kind of sad to see, for sure. That was another reason that I didn’t really go out very often, as well, because some places that I knew really well were closing down, and things like that. It was really sad. *(Kit, aged 20-24)* |
